# Effectiveness of the TYPHIBEV® (Vi-CRM197 conjugate) vaccine introduction in Nepal: a test-negative, case-control study

**DOI:** 10.1101/2025.11.20.25340644

**Authors:** Dipesh Tamrakar, Sabin Bikram Shah, Esther Jung, Shivram Naga, Basudha Shrestha, Pratibha Bista Roka, Rabin Sharma Pokharel, Ram Hari Chapagain, Akhil Tamrakhar, Manoj Mahoto, Surendra Kumar Madhup, Rajeev Shrestha, Kate Doyle, Isaac I. Bogoch, Stephen P. Luby, Denise O. Garrett, Wirongrong Cheirakul, Jason R. Andrews

## Abstract

**Introduction:** Typhoid fever, caused by *Salmonella* Typhi, remains a major cause of global morbidity and mortality. TYPHIBEV®, a Vi-CRM197 typhoid conjugate vaccine (TCV), received WHO prequalification based on immunogenicity data, but there have been no published data on clinical efficacy or effectiveness. We aimed to evaluate the effectiveness of TYPHIBEV®, which was introduced through a catch-up campaign and routine immunization in Nepal, in preventing blood culture-confirmed typhoid fever among children.

**Methods:** We conducted a test-negative, case-control study where typhoid test-positive cases were defined as vaccine-eligible pediatric patients who tested positive for *Salmonella* Typhi by blood culture at participating health facilities and test-negative controls were vaccine-eligible patients who tested negative for *S*. Typhi on blood cultures. We matched by age, location, date of blood culture, and surveillance site. We used conditional logistic regression to calculate odds ratios (OR), and vaccine effectiveness was calculated as 1-OR.

**Results:** Between October 1, 2022 and December 31, 2024, 40 typhoid cases and 113 matched, test-negative controls were enrolled. Both cases and controls were similar in sociodemographic characteristics and water, sanitation and hygiene-related living conditions. Among 39 cases and 108 controls with known vaccine status, 20 cases (51%) and 91 controls (84%) had received TCV. Vaccine effectiveness was 89% (95% CI: 65-97%) and was lower among children <5 years (72%, 95% CI: -203-97%) compared with those 5-15 years (98%, 95% CI: 80-100%). Vaccine effectiveness estimates did not significantly differ when restricted to participants with documented vaccination status.

**Conclusion:** The findings indicate that TYPHIBEV® was highly effective in preventing typhoid fever up to 30 months following a national introduction, with effectiveness estimates comparable to those observed for Typbar-TCV®.

## INTRODUCTION

Typhoid fever, caused by *Salmonella* Typhi, remains a major cause of global morbidity and mortality. The World Health Organization (WHO) estimates an annual global burden of 11-20 million typhoid cases and 128,000-161,000 deaths per year^1^. While many high-income countries have successfully eliminated typhoid through improvements to water and sanitation, typhoid continues to pose significant public health burden in many low-and middle-income countries like Nepal. The incidence of typhoid in Nepal exceeds the WHO-defined threshold of 100 cases per 100,000 population per year for classification as a high burden country^2,3^. Within Nepal, typhoid remains a problem in both rural and urban communities^4^. Despite the high burden of typhoid, control efforts have achieved limited success, hindered by inadequate water, sanitation and hygiene infrastructure^3,5^. These limitations underscore the urgent need for effective prevention strategies.

In recent years, advances in typhoid vaccine development have offered new opportunities for prevention. The WHO prequalified the first typhoid conjugate vaccine (TCV), Typbar-TCV^®^, manufactured by Bharat Biotech in 2017^6^. Three subsequent randomized trials, conducted in Nepal^7^, Malawi^8^, and Bangladesh^9^, demonstrated 79-85% efficacy in preventing typhoid fever. Additionally, real-world effectiveness studies of Typbar-TCV in both outbreak (Pakistan, Zimbabwe) and non-outbreak (Mumbai, India) settings found vaccine effectiveness ranging from 72% to 95%^10–13^.

Three other conjugate vaccines were subsequently prequalified based on immunogenicity data, but there have been no studies demonstrating their efficacy or effectiveness in disease prevention. In April 2022, Nepal introduced one of these, the TYPHIBEV^®^ (Vi-CRM197 conjugate) vaccine, through a nationwide catch-up campaign targeting children between the ages of 15 months and 15 years, followed by introduction into routine expanded program for immunization (EPI) for children at 15 months of age, administered alongside the second dose of the measles and rubella (MR) vaccine^14^. We leveraged this introduction to evaluate the effectiveness of TYPHIBEV® in preventing blood culture-confirmed typhoid fever among children in Nepal.

## METHODS

### Study sites

We recruited participants from seven health facilities located in and around the Kathmandu Valley: Bir Hospital, Dhulikhel Hospital Kathmandu University Hospital, Helping Hands Community Hospital, Siddhi Memorial Women and Children Hospital, Civil Service Hospital, Kanti Children’s Hospital, Kathmandu Model Hospital KMH. These hospitals were part of the Surveillance for Enteric Fever in Asia Project (SEAP) study, a hospital-based surveillance project for enteric fever^3^.

Five of the participating facilities (Bir Hospital, Kanti Children’s Hospital, Civil Service Hospital, Kathmandu Model Hospital, and Helping Hands Community Hospital) are located within Kathmandu, the capital and most populous city in Nepal. Kanti Hospital is the largest government-operated pediatric hospital in the country. Siddhi Hospital, a pediatric-only facility, is located in Bhaktapur, a city adjacent to Kathmandu. Dhulikhel Hospital Kathmandu University Hospital is located approximately 30 kilometers east of Kathmandu in the Kavrepalanchok (Kavre) District and serves a peri-urban and surrounding rural population.

### Study design

We conducted a test-negative, case-control study to evaluate the effectiveness of the vaccine in preventing symptomatic, blood culture-confirmed typhoid fever. We defined typhoid test-positive cases as vaccine-eligible pediatric patients who tested positive for *Salmonella* Typhi by blood culture at participating health facilities. Test-negative controls were vaccine-eligible patients who presented to the same facilities but tested negative for *S*. Typhi on blood culture.

### Inclusion criteria

We included children who were eligible for the typhoid vaccine catch-up campaign initiated in April 2022 (born between May 1, 2006 and January 1, 2021, corresponding to an age range of 15 months to 15 years and 11 months on April 1, 2022). We also included children eligible for typhoid vaccination under the routine immunization program starting on July 1, 2022 (children aged 15 months or older at the time of blood culture). A child within these age groups who tested positive for *Salmonella* Typhi in blood culture was classified as a case. Each case was matched to up to 3 controls from the same healthcare facility.

Controls were selected based on following criteria: 1) blood culture negative for *S*. Typhi, with preference, where possible, given to controls with laboratory-confirmed alternate etiologies; 2) blood culture performed within 28 days before or after the case’s culture date; 3) residence in the same geographic area as the case at the time of presentation and blood culture, categorized into five areas: Kathmandu, Bhaktapur, Lalitpur, urban areas in Kavre, rural areas in Kavre; and 4) age group matched to the case: 15 months to under 5 years, 5 to 9 years, or 10 to 15 years.

### Exclusion Criteria

Participants were excluded if they resided outside of the areas of Kathmandu, Bhaktapur, Lalitpur and Kavre; if they had lived in these areas for less than three months prior to presentation at the health facility in which their blood culture was collected; or, for individuals not eligible for routine immunization with TCV, if they were not present in Nepal during the catch-up campaign. Additional exclusion criteria included the lack of a valid phone number recorded at the health facility or if they declined to provide informed consent.

### Sample size

We estimated the requisite sample size to achieve 90% power for a matched case control study assuming vaccine effectiveness of 80%^15^, vaccine coverage of 90%, an average of three controls per case, and an exposure correlation of 0.30. With these assumptions, the estimated sample size was 44 cases and 132 controls. Sample size calculations were done with the “*epiR”* package in R (version 4.0.4).

### Data collection

The study team reviewed patient data on a weekly basis to identify potential cases based on age and diagnosis. Upon identification, study staff called the parents or guardians of patients with positive blood cultures for *S*. Typhi to assess interest and eligibility for study. Upon receiving permission, they visited the residence of these patients, where they obtained informed consent and administered sociodemographic questionnaires using the REDCap mobile app. Data was regularly uploaded to a password-protected central server hosted at Stanford University. The study team identified potential controls from laboratory blood culture records and SEAP surveillance data among test-negative participants who fit the matching criteria. We first prepared a sampling frame of potential controls matched by age group and time period. Whenever available, we purposefully selected controls with a blood culture-confirmed alternative etiology (i.e. a blood culture positive for other pathogenic bacteria). If alternate etiology controls were not available, we generated a random list of controls using a computer algorithm. As with the case recruitment, they called the parents or guardians of the potential controls to assess interest and eligibility for the study and visited the residence to administer questionnaires to those providing written informed consent. We recruited participants sequentially in descending order from the list until 3 controls were enrolled or the list was exhausted. The following information was collected with the questionnaires: gender, age, profession, place of residence, socioeconomic status, signs and symptoms associated with the current episode, contact with individuals with typhoid fever, type of water and food consumed, use of soap, toilets and latrines, and TCV vaccination status.

Vaccination status was verified with campaign-specific or routine immunization vaccination cards; immunization records in health facilities, government office (ward office), or schools (which kept records from the campaign); or by verbal ascertainment from parents. Research assistants recorded the date, type, and place of vaccination.

### Statistical analysis

We conducted conditional logistic regression analyses in R (version 4.0.4) using the *“survival”* package^16^ to compare the odds of vaccination among typhoid-confirmed cases and typhoid-negative controls. To calculate adjusted odds ratios (aORs) and adjusted vaccine effectiveness (VE), we included adjustments for continuous age and sex. Matched aORs of vaccination were used to calculate vaccine effectiveness (VE = [1 – aOR] × 100%).

In addition, we stratified analyses by age groups (15 months to <5 years vs. 5 to 15 years) to compare age-specific vaccine effectiveness. We published the statistical analysis plan prior to initiating data analysis (github.com/jasonandr/Typhibev-TND).

### Ethical considerations

We obtained written informed consent from parents and legal guardians and written assent from pediatric participants aged >7 years. The study protocol was approved by Nepal Health Research Council, Ethical Review Board (ERB: 395-2022); Kathmandu University School of Medical Sciences Institutional Review Committee KUSMS-IRC(IRC: 56/19); Kanti Children’s Hospital Institutional Review Committee (IRC:1535); Kathmandu Model Hospital Institutional Review Committee (IRC: 066-2022); and Stanford University Institutional Review Board (IRB:39627).

## RESULTS

A total of 108,678 blood cultures were performed for individuals presenting with fever or sepsis at any of the participating surveillance sites during the study period. Among these, 235 blood cultures were positive for *S.* Typhi. Among these individuals, 188 (80%) were outside of the eligible age group and 2 were outside catchment area.

Among the 45 individuals who were considered eligible cases according to the inclusion criteria, 40 (88%) consented to participate in the study: 11 (27.5%) were aged 15 months to <5 years, and 29 (72.5%) were aged 5 to 15 years. During the study period, total of 108,443 blood cultures were negative for *S*. Typhi. Of these, 50,711 were excluded because individuals were outside age eligible range (<15 months or >15 years), leaving 57,732 age-eligible individuals with typhoid-negative blood cultures to serve as controls. After matching for window of blood culture collection (±28 days), age group (15 months to <5 years, 5 to 9 years, 10 to 15 years), and area of residence (Kathmandu, Bhaktapur, Lalitpur, Kavre), we included 113 participants as controls in the analysis (**Figure 1**). There were 3 controls for 33 cases and 2 controls for 7 cases.

**Figure 1:**
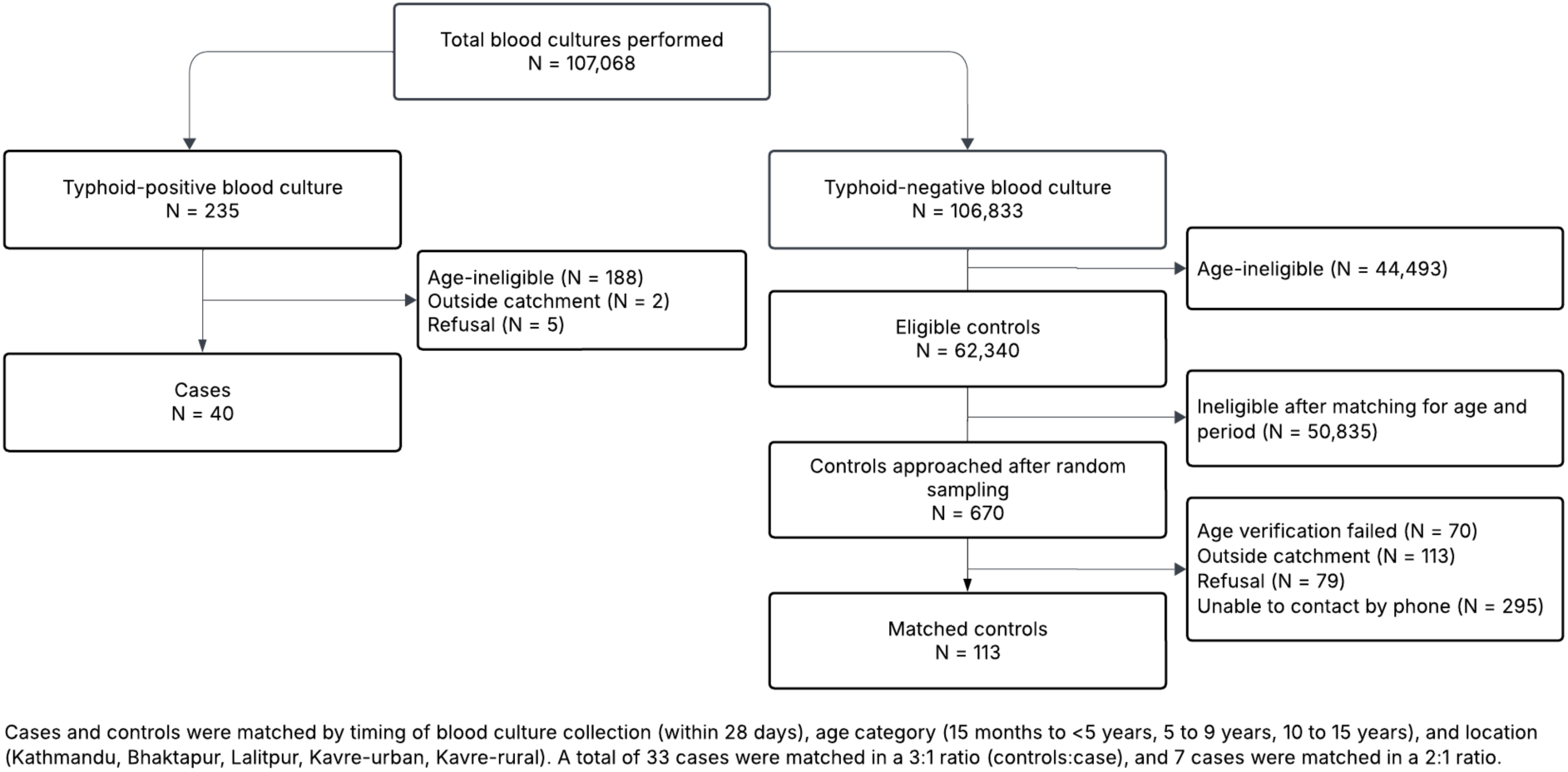
Flow diagram of case and control recruitment and matching.

When a case was positive, we prepared a list of potential controls from the blood culture database by matching the time period (plus minus 28 days) and age category. We purposefully selected alternate etiology controls first, if any, and randomly ordered the remaining potential controls. We approached these controls sequentially from the top to bottom of the randomized list. During the interview, we confirmed the matching criteria like location, age etc. We continued recruitment until we enrolled three controls per case. For individuals whose phone numbers were busy or switched off, we attempted to contact them up to three times before moving to the next potential control. There were 19 alternate etiology controls matched for 9 cases; the most common alternative etiologies were: *Staphylococcus aureus* (16/19), *Acinetobacter* spp. (2/19), and *E. coli* (1/19).

Cases and controls had similar sociodemographic characteristics (**Table 1**). Controls reported slightly better water, sanitation and hygiene (WASH) practices compared to cases; however, these differences were not statistically significant (**Table 2**). Among the 40 matched cases, 50% were vaccinated compared to 81% of the 113 matched controls. Among those vaccinated, confirmation of vaccine receipt in 60% of cases and 65% of controls was based on vaccine card or school or health records, while the remainder were based on parent or guardian recall (**Table 2**).

**Table 1.** Characteristics of typhoid cases and controls.

| Characteristic | Cases<br>N = 40 | Controls<br>N = 113 | P value* |
| --- | --- | --- | --- |
| Age categories |  |  |  |
| 15 months to <5 years | 11 (27.5) | 32 (28.3) |  |
| 5 years to 9 years | 11 (27.5) | 30 (26.5) |  |
| ≥10 years | 18 (45) | 51 (45.1) |  |
| Median age (IQR) | 9.1 (4.9-12.8) | 8.9 (4.0-12.8) |  |
| Gender |  |  | 0.847 |
| Male | 25 (62.5) | 71 (62.8) |  |
| Female | 15 (37.5) | 42 (37.2) |  |
| Ethnicity |  |  | 0.405 |
| Brahmin/Chettri | 15 (37.5) | 59 (52.2) |  |
| Janjati | 7 (17.5) | 21 (18.6) |  |
| Newars | 12 (30) | 19 (16.8) |  |
| Dalit | 3 (7.5) | 9 (8) |  |
| Madhesi | 3 (7.5) | 5 (4.4) |  |
| Father's Education |  |  | 0.902 |
| No formal education | 3 (7.5) | 6 (5.3) |  |
| Primary | 17 (42.5) | 49 (43.4) |  |
| Secondary | 12 (30) | 36 (31.9) |  |
| Post-secondary or more | 8 (20) | 22 (19.5) |  |
| Mother's Education |  |  | 0.077 |
| No formal education | 8 (20) | 12 (10.6) |  |
| Primary | 11 (27.5) | 51 (45.1) |  |
| Secondary | 12 (30) | 37 (32.7) |  |
| Post-secondary or more | 9 (22.5) | 13 (11.5) |  |
| Material of house |  |  | 0.546 |
| Cement with brick/stone | 37 (92.5) | 108 (95.6) |  |
| Mud with brick/stone | 1(2.5) | 3 (2.7) |  |
| Tin | 2 (5) | 2 (1.8) |  |

Data presented as n(%) unless specified otherwise.
\*Calculated using likelihood ratio tests from conditional logistic regression models accounting for the matched study design. P-values not reported for matching variables.

**Table 2.**
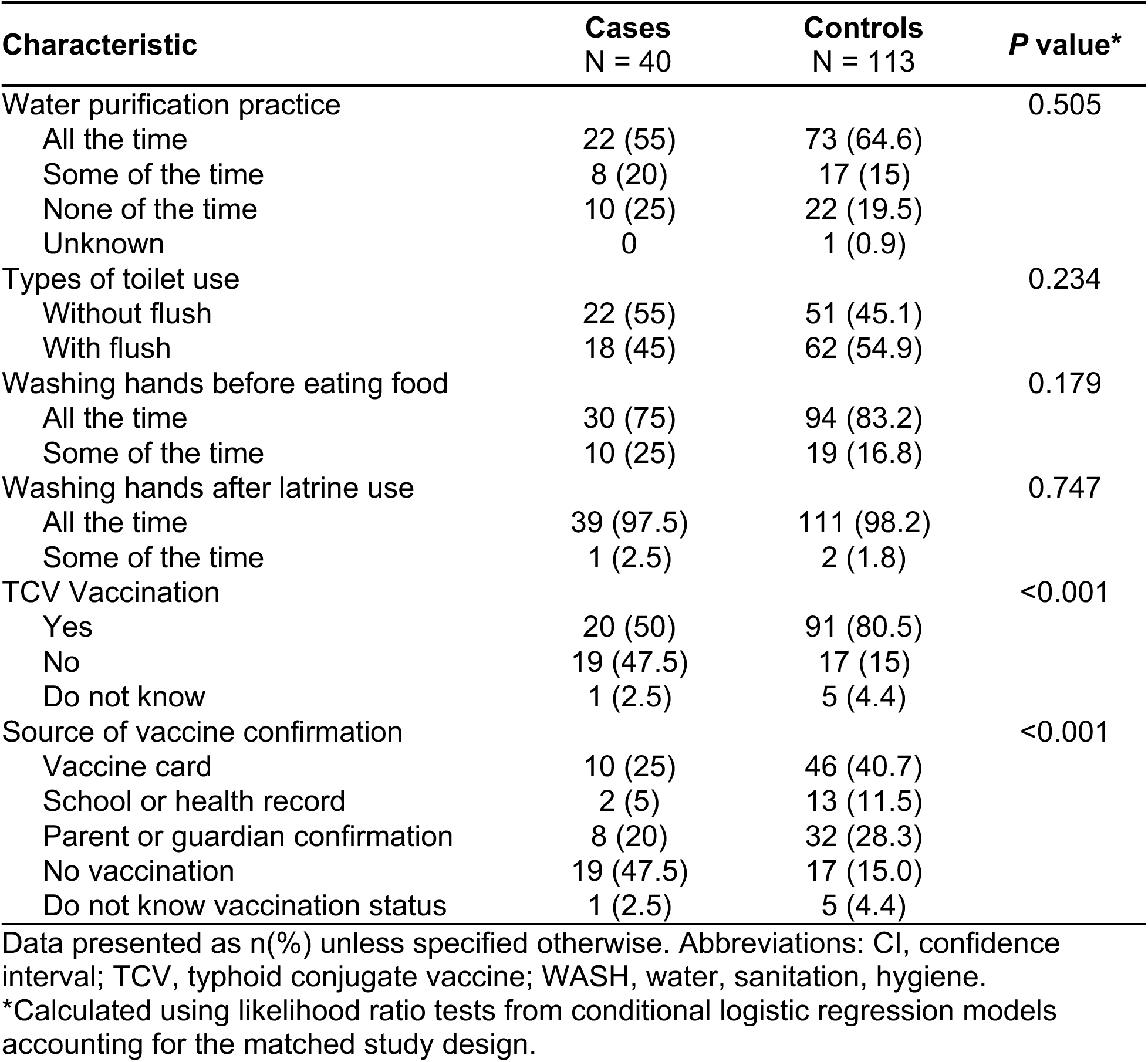
Comparison of WASH Practices and vaccination status among typhoid cases and controls.

| Characteristic | Cases<br>N = 40 | Controls<br>N = 113 | P value* |
| --- | --- | --- | --- |
| Water purification practice |  |  | 0.505 |
| All the time | 22 (55) | 73 (64.6) |  |
| Some of the time | 8 (20) | 17 (15) |  |
| None of the time | 10 (25) | 22 (19.5) |  |
| Unknown | 0 | 1 (0.9) |  |
| Types of toilet use |  |  | 0.234 |
| Without flush | 22 (55) | 51 (45.1) |  |
| With flush | 18 (45) | 62 (54.9) |  |
| Washing hands before eating food |  |  | 0.179 |
| All the time | 30 (75) | 94 (83.2) |  |
| Some of the time | 10 (25) | 19 (16.8) |  |
| Washing hands after latrine use |  |  | 0.747 |
| All the time | 39 (97.5) | 111 (98.2) |  |
| Some of the time | 1 (2.5) | 2 (1.8) |  |
| TCV Vaccination |  |  | <0.001 |
| Yes | 20 (50) | 91 (80.5) |  |
| No | 19 (47.5) | 17 (15) |  |
| Do not know | 1 (2.5) | 5 (4.4) |  |
| Source of vaccine confirmation |  |  | <0.001 |
| Vaccine card | 10 (25) | 46 (40.7) |  |
| School or health record | 2 (5) | 13 (11.5) |  |
| Parent or guardian confirmation | 8 (20) | 32 (28.3) |  |
| No vaccination | 19 (47.5) | 17 (15.0) |  |
| Do not know vaccination status | 1 (2.5) | 5 (4.4) |  |

Data presented as n(%) unless specified otherwise. Abbreviations: CI, confidence interval; TCV, typhoid conjugate vaccine; WASH, water, sanitation, hygiene.
\*Calculated using likelihood ratio tests from conditional logistic regression models accounting for the matched study design.

For the primary analysis, we excluded 6 individuals (1 case and 5 controls) who were not sure about their vaccination status. Among the remaining participants, vaccinated individuals had 89% lower adjusted odds of testing positive for typhoid on blood culture compared to unvaccinated individuals when adjusting for age and sex (adjusted odds ratio [aOR] 0.11; 95% confidence interval [CI]: 0.03–0.35), corresponding to a VE of 89% (95% CI: 65-97%). When we restricted the definition of vaccination to those confirmed by vaccine card or government records, vaccination was associated with a similar 88% reduction in the odds of testing positive for typhoid (aOR 0.12; 95% CI: 0.03 - 0.47) (**Figure 2**). In sensitivity analysis, when individuals with unknown vaccination status were classified as unvaccinated, vaccine effectiveness remained stable at 89% (95% CI: 57-97%).

**Figure 2.**
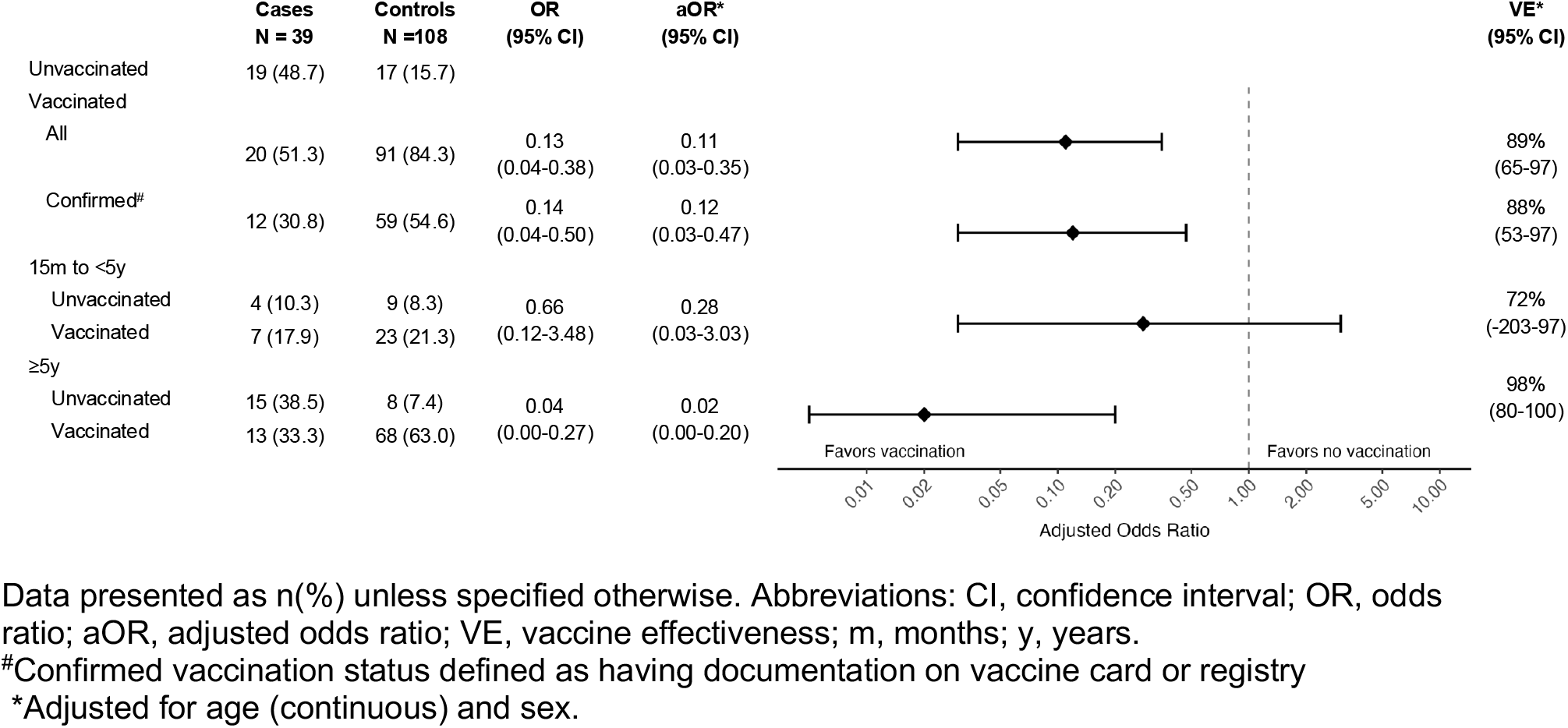
Conditional logistic regression and vaccine effectiveness of vaccination among cases and controls, Nepal, 2022-2024.

Among participants in the younger age group (15 months to <5 years), vaccination was associated with a 72% reduction in odds of testing positive (aOR 0.28; 95% CI: 0.03–3.03), although this effect was not significant. Among participants 5 years and older, vaccination was associated with a 98% reduction in odds of testing positive for typhoid (aOR 0.02; 95% CI: 0.00–0.20), corresponding to a VE of 98% (95% CI: 80-100%) (**Figure 2**). There was a statistically significant difference in vaccine effectiveness estimates between the age groups (p = 0.026), suggesting greater vaccine-mediated protection among children 5 years and older.

## DISCUSSION

This study provides the first evidence of the effectiveness of the TYPHIBEV® typhoid conjugate vaccine when implemented through a nationwide catch-up campaign and subsequent introduction into the routine immunization program in Nepal. Among children between 15 months and 15 years of age, we found that TYPHIBEV® TCV was 89% effective in preventing typhoid fever. Vaccine effectiveness was lower among children under 5 years of age compared to those of ages 5-15 years. Together, these findings indicate that TYPHIBHEV® was highly effective in typhoid prevention among children in Nepal.

The vaccine effectiveness of TYPHIBEV® in the current study was similar to estimates of protection conferred by Typbar-TCV in clinical trials (79-85%)^7–9^ and post-introduction observational studies (72-95%)^10–13^. Variability in VE estimates across studies may be attributed to differences in the force of infection, prior population immunity, timing of vaccination relative to exposure, and study design.

We found lower vaccine effectiveness in children aged 15 months to <5 years (72%), compared to children aged 5 to 15 years (98%). A similar pattern was observed in the vaccine effectiveness study conducted in Navi Mumbai, where effectiveness was 72.3% in children under 5 years and 87.0% in those aged 5 to 14 years^11^. This age-related trend has also been reported in a recent meta-analysis of efficacy studies, which estimated pooled efficacy at 73% for children under 5 years and 87% for those over 5 years of age^17^. The observed trend of lower vaccine effectiveness in younger children is thought to result from their less mature immune systems, where children are able to mount T-cell–dependent responses, but their immune memory and antibody affinity maturation are still developing^18–20^. This may lead to weaker initial immune responses and a more rapid decline in protective antibody levels over time, whereas older children tend to mount stronger and more durable immune responses^17^.

Recent studies have found considerable waning of Typbar-TCV efficacy over time. Extended follow-up of the randomized trial in Bangladesh demonstrated that vaccine efficacy declined from 84% in those vaccinated within the past three years compared to 55% among those vaccinated 3-5 years earlier^21^. In contrast, efficacy did not wane rapidly in extended follow up from the clinical trial in Malawi, declining modestly from 83% after one year to 78% after 4.6 years^8^.

The results of this study should be interpreted within the context of several limitations. Given the moderate sensitivity of blood culture (approximately 60%)^22^, some true typhoid cases may have been misclassified as controls due to false-negative results, potentially leading to an underestimation of vaccine effectiveness. Although we prioritized the selection of controls with alternative etiologies to minimize this risk, and the negative predictive value of a negative blood culture for typhoid in Nepal exceeds 90%^23^, some degree of misclassification is expected. A recent study comparing the test-negative design with the cohort-based analysis of clinical trial data confirmed the validity of the former approach^24^. Additionally, about one-fourth of participants self-reported their vaccination status, introducing possible recall bias. However, vaccine effectiveness estimates remained robust in sensitivity analyses restricted to cases with documented vaccination, suggesting that the potential bias had minimal impact. Finally, the introduction of TCV occurred after the COVID-19 pandemic, during a period of reduced typhoid transmission^25^, which may have inflated the observed vaccine effectiveness due to a lower force of infection.

The study provides the first real-world evidence of the effectiveness of TYPHIBEV®, the WHO-prequalified typhoid conjugate vaccine, in a mass vaccination campaign and routine immunization program. The findings indicate that TYPHIBEV® is highly effective in preventing typhoid fever, with effectiveness estimates comparable to those observed for Typbar-TCV in similar settings. These results reinforce the utility of typhoid conjugate vaccines as a critical component of typhoid control strategies in endemic countries and provide valuable data to guide national and global policy decisions around vaccination.

## Data Availability

We published the statistical
analysis plan prior to initiating data analysis.

https://github.com/jasonandr/Typhibev-TND

## Acknowledgements

The study team is grateful for the time and effort of the many hospital staff who enrolled and followed up patients. The patient participants are the reason this study was possible—we would not have these findings to report without their generous contributions.

